# Optimizing mixed sample analysis as a step to comprehensive desease screening: a pilot study

**DOI:** 10.1101/2023.11.07.23297590

**Authors:** Lucia Krasnicanova, Natalia Forgacova, Tatiana Sedlackova, Jaroslav Budis, Juraj Gazdarica, Vanda Repiska, Tomas Szemes

## Abstract

**Background:** Lynch Syndrome (LS) is an autosomal dominant hereditary syndrome associated with a diverse range of cancer types. Despite being one of the most prevalent hereditary cancer syndromes, the detection of LS remains challenging due to the absence of well-defined diagnostic criteria which would be able to select all patients who should undergo testing for LS and the limitations of existing screening methods. The implementation of an efficient screening program capable of accurately detecting the majority of LS cases remains a topic of continuous discussion in the scientific literature, with recent studies emphasizing the significance of a universal screening program.

**Methods:** Our study aimed to develop and optimize a cost-effective universal screening method for detecting mutation in the mismatch repair (MMR) genes through mixed sample analysis. We tested five approaches in terms of the use of biological material and the analysis of mixed samples.

**Results:** Each approach successfully detected a specific Lynch-associated pathogenic variant in mixed in the pooled samples with frequency 5.00%, with the lowest allelic fraction recorded at 3.04%. Approach 2, which involved isolating DNA from each patient individually, demonstrated the highest average allelic fraction (7.04%). However, considering financial and time requirements, approach 1, where DNA was isolated only after mixing aliquots of whole blood, proved to be the most favorable.

**Conclusion:** The findings of our study present a promising opportunity to improve LS detection. The identification of LS not only has the potential to prevent cancer-related morbidity and mortality but also facilitates continued progress in understanding the primary prevention of cancer.

## Introduction

Lynch syndrome (LS) is an autosomal dominant hereditary syndrome caused by germline mutations in the DNA mismatch repair (MMR) genes *MLH1, MSH2, MSH6, PMS2*, and/or in the Lynch-associated gene *EPCAM* [1,2]. LS is among the most common hereditary cancer syndromes, with an estimated population frequency of 0.36 % (1 in 279 people) [3]. Individuals with LS have an increased risk of developing a specific cancer type, especially colorectal cancer (CRC) and endometrial cancer (EC), as well as gastric, ovarian, pancreatic, prostate, urinary tract, kidney, small intestine, hepatobiliary tract, brain, and cutaneous cancer. The association of LS with breast cancer, as suggested by some studies, remains questionable [4].

The main problem in detecting LS is the fact that many cases remain undetected due to the diverse range of associated cancer types and the absence of well-defined diagnostic criteria. These criteria undergo modifications in their models and strategies over time due to the recognition of their inadequacy based on practical experience [5]. Initially, individuals eligible for genetic testing for LS were identified based on their fulfillment of the Amsterdam criteria or Bethesda guidelines [6]. However, it has been established that these criteria are insufficient for effectively screening for Lynch syndrome because they can miss 23-50% of cases [3,7,8]. Adar et al. [9] observed that no clinical criteria can accurately identify all LS patients detected through universal germline mutation screening. Furthermore, Moreira et al. [10] also highlighted the limitations of the concurrent criteria and methods. Among 10,019 patients analyzed, only 85 (27.2%) fulfilled the Amsterdam criteria I and II, and merely 214 (68.6%) met at least one criterion within the revised Bethesda criteria among the identified 312 LS patients. Additionally, predictive computer models such as PREMM5 have demonstrated ineffectiveness in clinical practice, primarily due to their focus on identifying individuals at the highest risk rather than encompassing the entire spectrum of LS patients [10,11]. As a consequence of these limitations, only a fraction of LS cases undergo genetic testing.

The diagnosis of LS involves two standard laboratory tests, DNA microsatellite instability (MSI) analysis and immunohistochemistry (IHC) of MMR proteins. A definitive diagnosis of LS must be confirmed by detecting the germline mutations in MMR genes [12,13]. The current strategy for LS detection involves universal screening of Lynch-associated tumors using these methods [14]. While tumor testing for MSI is sensitive, it is not specific enough for LS, as only 20-25% of MSI-H tumors are associated with a germline MMR gene mutation. Additionally, MSI testing fails to detect LS in approximately 5% of mutation carriers and has a sensitivity of only 86% for CRC in families with an MSH6 gene mutation. To complement MSI testing, immunohistochemistry screening is recommended [15]. Although PCR methods are currently the gold standard for determining MSI status, NGS-based methods are being developed for MSI analysis in various cancer types [16]. MSI NGS offers advantages such as analyzing a larger number of loci, reducing the threshold for identifying MSI-H tumors to 20%. Moreover, it demonstrates high sensitivity (96-100%) and specificity (97-100%) for LS screening [11]. With the decreasing costs of NGS, it may become more cost-effective to sequence more genes rather than relying on MSI analysis and immunohistochemistry [16,17].

The main aim of our study was to develop and optimize a cost-effective screening method for detecting mutations in the MMR genes through different approaches of mixed sample analysis. To test and evaluate the effectiveness of our method, we mixed an LS patient sample into the sample pool of a healthy population to detect a known mutation and quantify its proportion within the mixed sample. The analysis of mixed samples using this approach holds particular significance, particularly in LS-related cancers where LS is relatively uncommon. The findings of our study indicate a promising opportunity to enhance the effectiveness of universal screening for LS as it would be possible to simultaneously analyze samples from 10 patients by performing a single analysis. This improvement has the potential to greatly contribute to the development of more targeted therapeutic approaches for patients.

## Material and Methods

### Characterization of sample set

In order to make screening more cost-effective, we aimed to create and refine a method that could effectively detect mutations in the MMR gene in mixed patient samples. To test our method, we intentionally mixed one sample from a patient with Lynch syndrome with another nine control samples without the causal mutation into the sample pool to see if we could detect a known mutation and properly determine the fraction of that mutation in the sample. Our optimization efforts focused on pooling samples at various stages of sample processing workflow using a commercial kit for NGS library preparation.

### Types of sample pooling procedures

To find out the best way to mix the samples, we tested the following five different processing methods and separately prepared samples as controls. The first method, WB-pre-Iso, involved mixing of equivalent volume whole blood samples from individual probands before DNA isolation. In the second method, WB-post-Iso, samples were mixed after DNA isolation from whole blood of individual probands, with normalization to 10 ng/μl from each sample. The third protocol, BC-pre-Iso, used a mixture of buffy coats from individual probands for DNA isolation. Alternatively, DNA samples isolated from buffy coats of individual probands were normalized to 10 ng/μl and then pooled. Another method involved mixing the samples after the purification step that follows the multiplex PCR reaction for target amplification in the NGS library preparation process. In this case, the DNA samples were isolated from the buffy coat. Finally, separately prepared libraries from DNA samples isolated from the buffy coat of each proband were used as controls. All tested methods except the control preparation were performed in triplicate (Figure 1).

**Figure 1.**
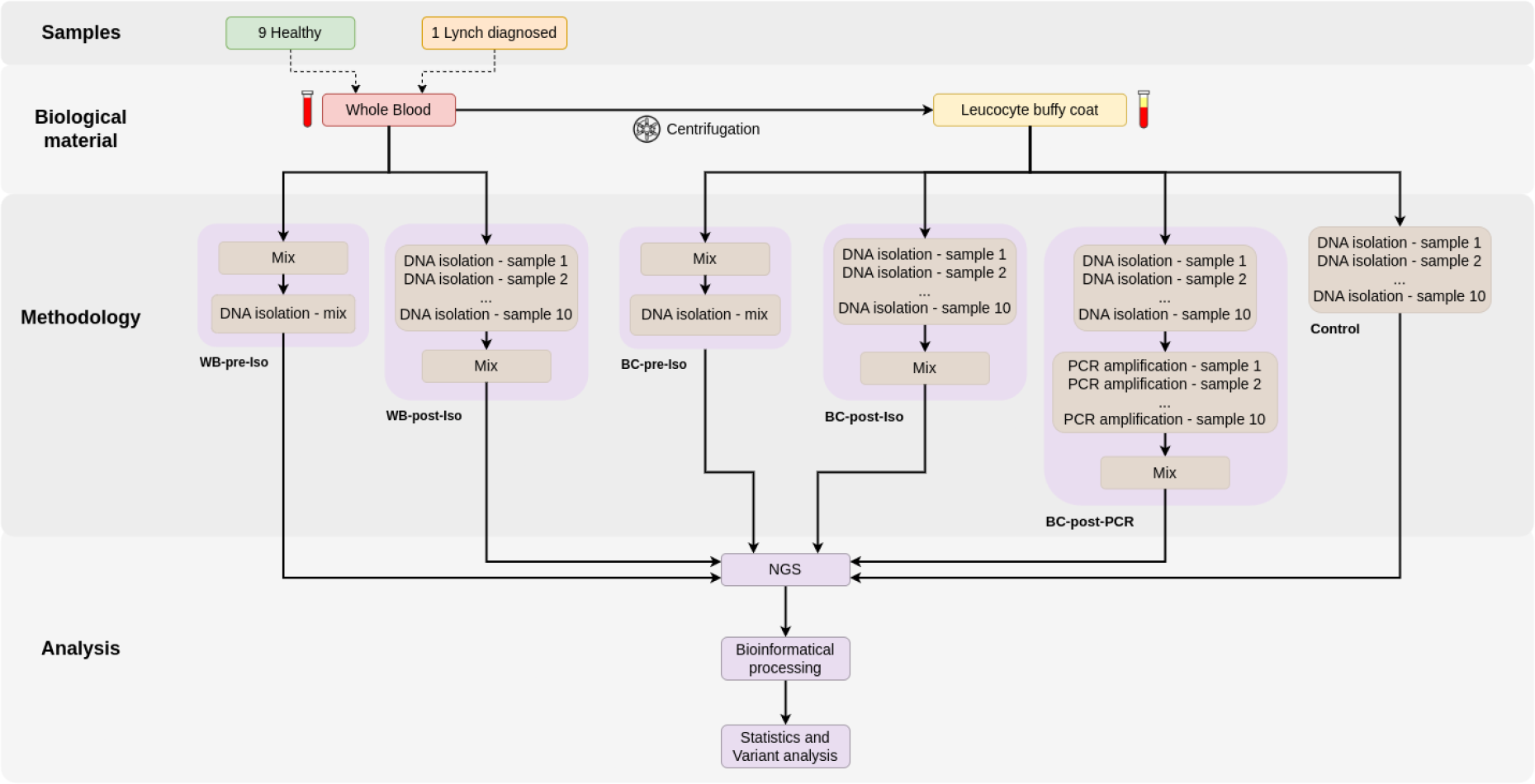
The diagram represents the comprehensive progression of the study, beginning from detailing of sample types, transitioning to the collection of materials, and finally branching out to various methodological approaches for analyzing grouped samples. Different methodology approaches are described as follows: WB-pre-Iso = Mix of Whole Blood before DNA isolation and sequencing; WB-post-Iso = Mix of Whole Blood after DNA isolation and before sequencing; BC-pre-Iso = Mix of Buffy Coats before DNA isolation and sequencing; BC-post-Iso = Mix of Buffy Coats after DNA isolation and before sequencing; BC-post-PCR = Mix of samples (DNA isolated from Buffy Coats) after PCR amplification and before sequencing; Control = Isolation from Buffy Coat of each participant.

### Sample processing

Venous blood from the probands was collected in BD Vacutainer blood collection tubes with the anticoagulant K2EDTA (BD, Plymouth, UK). The buffy coat was obtained by centrifuging the blood at 1000 g for 8 minutes, after which the plasma was removed, and the buffy coat was collected in a new tube. DNA isolations from both whole blood and buffy coat were performed using the QIAamp DNA Blood Mini Kit (Qiagen, Hilden, Germany) following the manufacturer’s protocol for spin isolation with some modifications. A recommended spin step, followed by incubating the spin column at 56°C for 2 minutes, was implemented to eliminate the chance of possible Buffer AW2 carryover. The isolated DNA samples were then eluted with 50 μl of millipore water and incubated for 10 minutes before undergoing a final centrifugation at 20,000 g for 1 minute. Concentrations of isolated DNA samples were determined fluorimetrically using Qubit 3.0, and samples were subsequently normalized to 10 ng/μl.

### NGS library preparation

A commercial kit, CleanPlex® Lynch Syndrome Panel (Paragon Genomics, Hayward, CA, USA), was used to prepare the NGS libraries, with 30 ng of DNA used as input. The libraries were prepared with half volume of reagents compared to the original protocol, where 10 cycles were used for target amplification during the multiplex PCR (mPCR) reaction. The second PCR reaction, used to amplify and index the libraries, was performed using i7 and i5 indexed primers suitable for sequencing by Illumina platforms. Six PCR cycles were used for library amplification. Final NGS libraries were quantified using Qubit 3.0 (Life Technologies, Singapore), and their length profile was determined based on on-chip electrophoresis using Agilent 2100 Bioanalyzer (Agilent Technologies, Waldbronn, Germany).

### NGS sequencing

Final NGS libraries were normalized to 2 nM and pooled equimolar. Sequencing was performed by Illumina MiSeq platform using MiSeq Reagent Kit v3 (Illumina, San Diego, CA, USA) as paired-end sequencing with reads length 151 bp.

### Bioinformatical processing

Adapters and low quality ends (<20) of sequenced reads were removed using Trimmomatic (v0.36; [18]), based on quality control statistics generated by FastQC (v0.11.5.). In the mapping step against the human genome (hg38), we employed BWA (v0.7.17.) to map the reads to the reference genome and SAMtools (v1.13; [19]) to sort and index generated SAM/BAM files. Finally, BCFtools (v1.13; [20]) with default settings was used for variant calling and normalization. All computational analyses were written and executed using the SnakeLines framework [21].

### Statistics and variant analysis

We analyzed Variant Call Format (VCF) files to obtain allele fractions (AF) for each analyzed sample. To achieve this, an in-house Python script was developed, utilizing the pyvcf (v0.6.8) library to analyze VCF files. For each variant in all sample VCF files, the script extracted the read depth (DP total) and the number of alt-forward and alt-reverse bases (DP alt), from which the alternative allele fraction (AF alt) was calculated as the ratio between DP total and DP alt. The allele count (AC alt) was also calculated as the sum of genotypes, where, for instance, a 0/1 genotype would result in an AC alt of 1. Subsequently, a resulting table was generated, containing only variants with an AC alt greater than 1 in at least one patient.

The specified metrics were evaluated for each set of three samples in all methodologies, which included WB-pre-Iso, WB-post-Iso, BC-pre-Iso, BC-post-Iso, BC-pre-PCR as well as 9 Control samples. The Control group served as a reference point to evaluate if the appropriate allele fraction was established in each experimental run. We also wanted to make sure that the analyzed mutation does not also occur in other members within the group we analyzed, which could affect the allelic fraction. Finally, the allele fraction was compared among the different methodologies to determine any notable differences. The target allele fraction for the observed variant was set to a value of 5.00%, owing to the fact that the mixture comprised samples from 10 individuals, out of which one harbored a mutation in one of their alleles.

## Results

Table 1. presents the outcomes of measuring the concentration of the final DNA library samples using Qubit 2.0 through fluorometric quantification. The data reveals that when sample aliquots were mixed before DNA isolation, lower DNA concentrations were observed. Among the different sample mixing approaches, the BC-post-Iso method, which involved mixing DNA samples from the leukocyte-thrombocyte fraction, exhibited the highest DNA concentration values. These results pertain to the DNA concentration values in the final library of the CleanPlex® Lynch Syndrome Panel kit, as determined by Qubit 2.0.

**Table 1.** The table depicts results from measuring the concentration of final DNA library samples using the Qubit 2.0 system. This system uses a fluorometric approach for more accurate quantification. Concentration was determined individually for each sample replicate to ensure robust data validity. *WB-pre-Iso* = Mix of Whole Blood before isolation and sequencing; *WB-post-Iso* = Mix of Whole Blood after isolation and sequencing; *BC-pre-Iso* = Mix of Buffy Coats before isolation and sequencing; *BC-post-Iso* = Mix of Buffy Coats after isolation and sequencing; *BC-post-PCR* = Mix of Buffy Coats after PCR amplification and sequencing; *Control* = Isolation from Buffy Coat of each participant.

| Sample | Concentration of final DNA library (ng/μl) |
| --- | --- |
| WB-pre-Iso 1 | 0,588 |
| WB-pre-Iso 2 | 0,886 |
| WB-pre-Iso 3 | 0,470 |
| WB-post-Iso 1 | 0,830 |
| WB-post-Iso 2 | 0,586 |
| WB-post-Iso 3 | 0,780 |
| BC-pre-Iso 1 | 0,278 |
| BC-pre-Iso 2 | 0,598 |
| BC-pre-Iso 3 | 0,760 |
| BC-post-Iso 1 | 1,220 |
| BC-post-Iso 2 | 1,110 |
| BC-post-Iso 3 | 1,310 |
| BC-post-PCR 1 | 0,570 |
| BC-post-PCR 2 | 0,690 |
| BC-post-PCR 3 | 0,736 |
| Control 1 | 1,600 |
| Control 2 | 1,810 |
| Control 3 | 1,440 |
| Control 4 | 2,040 |
| Control 5 | 1,380 |
| Control 6 | 1,360 |
| Control 7 | 1,110 |
| Control 8 | 1,910 |
| Control 9 | 1,690 |
| Control 10 | 1,490 |

Overall, it can be inferred that the quality of results from the CleanPlex® Lynch Syndrome Panel (Paragon Genomics) DNA library, as assessed by the Agilent 2100 Bioanalyzer, is better when a mixed sample of DNA from 10 patients is processed compared to when samples are purified separately and mixed after the first PCR step (see Supplementary figures 1, 2, 3 and Table 1). Additionally, consistent levels of amplification are observed in the control samples across all 10 patients (see Supplementary figure 4).

After sequencing and bioinformatics analysis of the samples, we then analyzed the allele fraction of the specific LS patient mutation within each mixed sample. In total, we were able to map 110 272 reads to the region across all samples. Of these, 5 541 contained a specific mutation (single nucleotide deletion C) located in the MSH6 gene on chromosome 2 at position 47 403 369 when mapping to hg38. The average coverage of that region per sample was 4 410.88 reads per sample. On average, the allele fraction of the mutation analyzed by us was 5.025%. Table 2 shows allele fraction, depth of coverage of the specified region, number of reads containing the mutation and total mapped reads for each sequenced sample. The fragments of the mixed samples containing the mutation accounted for 3.04% to 8.3% of the fragments mapped to a particular region.

**Table 2.** Allele fraction of the mutation located in the MSH6 gene at chromosome 2 position 47403369 (hg38), total depth of coverage of the region, number of reads containing the mutation for different mixing approaches and control samples. The table also provides data on the average depth of coverage and the standard deviation of depth of coverage for all variants with an AC > 1 in at least one mixing approach or control sample. *WB-pre-Iso* = Mix of Whole Blood before isolation and sequencing; *WB-post-Iso* = Mix of Whole Blood after isolation and sequencing; *BC-pre-Iso* = Mix of Buffy Coats before isolation and sequencing; *BC-post-Iso* = Mix of Buffy Coats after isolation and sequencing; *BC-post-PCR* = Mix of Buffy Coats after PCR amplification and sequencing; *Control* = Isolation from Buffy Coat of each participant.

| Sample | Allele fraction (%) | Coverage depth | Number of reads containing mutation | Average coverage depth (variants with AC > 0) | Standard deviation of coverage depth (variants with AC > 0) |
| --- | --- | --- | --- | --- | --- |
| WB-pre-Iso 1 | 3.68 | 6637 | 244 | 4114.4 | 2456.7 |
| WB-pre-Iso 2 | 3.18 | 4438 | 141 | 3202.2 | 2014.3 |
| WB-pre-Iso 3 | 4.19 | 3398 | 142 | 2539.8 | 1690.0 |
| WB-post-Iso 1 | 6.31 | 4646 | 293 | 3224.5 | 2033.4 |
| WB-post-Iso 2 | 8.30 | 3059 | 254 | 2338.2 | 1568.4 |
| WB-post-Iso 3 | 6.96 | 6320 | 440 | 4012.7 | 2399.0 |
| BC-pre-Iso 1 | 4.03 | 4514 | 182 | 3121.4 | 1967.4 |
| BC-pre-Iso 2 | 4.24 | 4123 | 175 | 3015.4 | 1886.2 |
| BC-pre-Iso 3 | 4.63 | 3974 | 184 | 2819.5 | 1807.7 |
| BC-post-Iso 1 | 5.56 | 4391 | 244 | 3116.8 | 1957.5 |
| BC-post-Iso 2 | 6.02 | 4933 | 297 | 3359.0 | 2028.9 |
| BC-post-Iso 3 | 6.37 | 3674 | 234 | 2599.5 | 1665.6 |
| BC-post-PCR 1 | 5.68 | 2746 | 156 | 2387.6 | 1672.1 |
| BC-post-PCR 2 | 4.08 | 2793 | 114 | 2334.8 | 1660.6 |
| BC-post-PCR 3 | 3.04 | 2825 | 86 | 2342.7 | 1627.8 |
| Control 1 | 0 | 6528 | 0 | 4171.6 | 2439.0 |
| Control 2 | 0 | 4265 | 0 | 3280.9 | 2075.6 |
| Control 3 | 0 | 4592 | 0 | 3312.5 | 2074.3 |
| Control 4 | 0 | 4377 | 0 | 3299.9 | 2040.4 |
| Control 5 | 48.90 | 4816 | 2355 | 3394.6 | 2041.6 |
| Control 6 | 0 | 4731 | 0 | 3139.4 | 2571.3 |
| Control 7 | 0 | 4189 | 0 | 3216.8 | 2075.6 |
| Control 8 | 0 | 3637 | 0 | 2947.5 | 1959.8 |
| Control 9 | 0 | 3580 | 0 | 2583.2 | 1674.8 |
| Control 10 | 0 | 4591 | 0 | 3448.5 | 2259.5 |

With approach 1, where DNA was isolated only after mixing aliquots of whole blood, we found an average coverage per sample of 4 824.3. The average number of reads containing a given mutation was 175.66. The allele fraction averaged for a given approach in the samples was 3.641%. For approach 2, where DNA from whole blood samples was first isolated and then mixed, the average number of reads covering the selected position was 4 675.0. The average number of reads where we detected the mutation was 329, resulting in an average allele fraction as high as 7.037%. In approach 3, where we mixed the buffy coats of the samples analyzed by us prior to DNA isolation, we observed an average read coverage of the selected region of 4 203.7 reads. On average per sample, the mutation was present in 180.33 reads, resulting in an average allele fraction of 4.29%. In approach 4, where we mixed the DNA from the buffy coat after isolation from each sample independently and then analyzed as a mixture sample, we recorded an average coverage of the selected site in the genome at 4 332.7 reads, with an average number of reads containing the mutation of 258.33 reads. From this, we determined an average allele fraction of 5.973%. The final approach in the mixed sample analysis was approach 5, where we amplified the samples after DNA isolation in the first PCR kit CleanPlex® Lynch Syndrome Panel (Paragon Genomics), and then after purification, we mixed and analyzed the samples as a single sample. The average coverage of the selected position was 2 788.0 reads. The mutation we analyzed was present in an average of 118.66 reads, which means that the average allele fraction was 4.256% (Table 3).

**Table 3.**
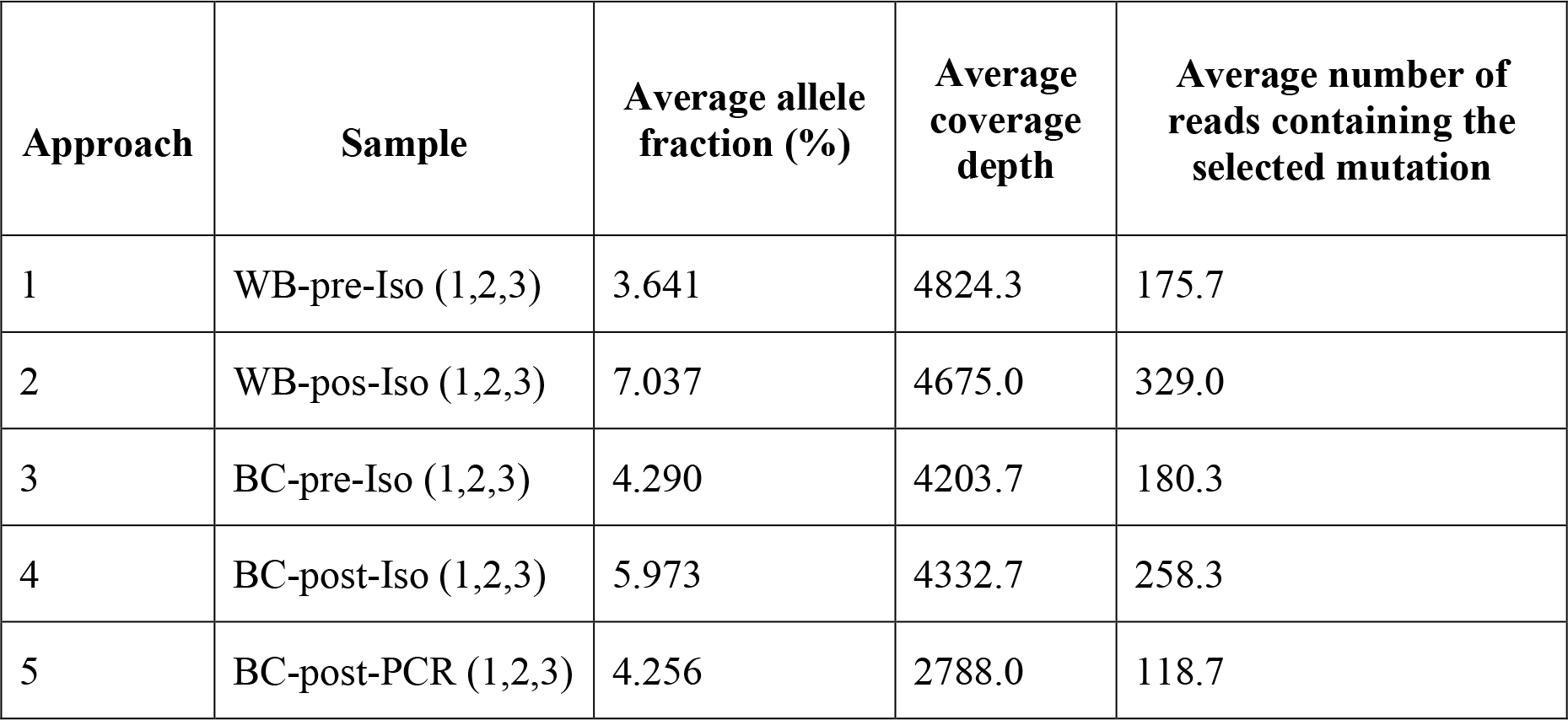
Averages of number of reads containing the selected mutation, coverage depth and allele fraction across different mixed sample analysis approaches.

| **Approach** | **Sample** | **Average allele fraction (%)** | **Average coverage depth** | **Average number of reads containing the selected mutation** |
| --- | --- | --- | --- | --- |
| 1 | WB-pre-Iso (1,2,3) | 3.641 | 4824.3 | 175.7 |
| 2 | WB-pos-Iso (1,2,3) | 7.037 | 4675.0 | 329.0 |
| 3 | BC-pre-Iso (1,2,3) | 4.290 | 4203.7 | 180.3 |
| 4 | BC-post-Iso (1,2,3) | 5.973 | 4332.7 | 258.3 |
| 5 | BC-post-PCR (1,2,3) | 4.256 | 2788.0 | 118.7 |

## Discussion

The established gold standard for LS diagnosis involves the identification of a hereditary mutation in any of the four MMR genes and in the non-MMR gene, *EPCAM*. However, the implementation of an efficient screening program capable of accurately detecting the majority of LS cases remains a topic of continuous discussion in the scientific literature. While some groups advocate for selective screening, recent studies strongly emphasize the significance of a universal screening program using genetic testing, encompassing all patients with Lynch-associated cancers, irrespective of clinicopathological characteristics or family history [22,23]. Our study aims to assess the cost- and time-effectiveness of different diagnostic strategies for LS from the perspective of mixed sample analysis. We intentionally combined a sample from a confirmed LS patient with samples from the healthy population, with the aim of identifying a sufficiently large percentage of DNA containing a mutation. We proposed different approaches in terms of the use of biological material and tested several options for the analysis of mixed samples, while the use of the leukocyte-platelet fraction after centrifugation of a unit of anticoagulated whole blood proved to be the most appropriate using the commercial kit CleanPlex® Lynch Syndrome Panel (Paragon Genomics).

During the optimization of mixed sample analysis, we conducted tests using various approaches (Table 3). Approach 1, where DNA was isolated only after mixing aliquots of whole blood, showed the lowest allelic fraction (3.641). This deviation from the expected allelic fraction could be attributed to variations in the leukocyte content among the patients’ blood samples, given that DNA isolation occurred after the samples were mixed. Therefore, we assumed that the results would be more accurate in the case of buffy coat collection. We confirmed this by observing a higher allelic fraction in approaches 3, 4 and 5 where we used a buffy coat for analysis. We reached the second lowest allelic fraction in approach 5, when the samples were mixed only after the first amplification step. Given that in this approach the samples were amplified separately in the first PCR reaction, it is the least financially and time-effective approach, as well as the least effective in terms of the potential of using the given screening method for a larger number of patients in one mixed sample. Approach 2 showed the highest average allelic fraction (7.037). This method involved isolating DNA from whole blood of each patient individually, making it less advantageous in terms of time and cost compared to approaches 1 and 3, which utilized mixed samples from whole blood or buffy coats. From a statistical standpoint, approaches 3 and 4 yielded results closest to the expected allelic fraction of approximately 0.05. Overall, each approach successfully detected a specific pathogenic variant in patient with Lynch syndrome, with the lowest allelic fraction of 0.0304 in the case of sample BC-post-PCR 3.

In recent years, there have been rapid developments in the identification and management of individuals and families affected by Lynch syndrome. Advances in molecular testing and NGS technologies have greatly improved the ability to screen all patients not only with CRC but also with other Lynch-related cancers. According to many studies, mentioned in the introduction, testing all patients with Lynch associated cancer would increase the number of diagnosed patients. Considering that if a mutation causing LS occurs in a mixed sample, it is necessary to test each patient separately, the method proposed by us would be beneficial especially in Lynch-associated cancers, where patients with LS do not occur statistically as often as in colorectal and endometrial cancer. Screening using mixed samples could be very beneficial, considering the number of tested patients without capturing a patient with LS and thus without the need to test every individual from a pool of patients within a mixed sample. However, we can also expect a reduction in screening costs for patients with colon and endometrial cancer, given that LS causes statistically 2 to 4% of colorectal cancers and approximately 2.5% of endometrial cancers [24].

Although our study showed promising results, there are some limitations that need to be taken into account in the further interpretation of the results. The study may be limited by the sample size, as it might not fully represent the diversity of LS cases in larger populations. However, it is important to point out that the goal of this study was to find the most suitable way of processing the sample with subsequent application in population study in the future. While our results demonstrated the successful detection of the LS pathogenic variant, the allelic fraction recorded in some cases might still be below the threshold for reliable clinical diagnosis, indicating potential limitations in sensitivity. Although many approaches in the study have been shown to be cost and time-effective, it’s crucial to consider the practicality and resources required for implementation in a real clinical setting. In summary, the study provides promising results, but further research and validation in larger and more diverse patient cohorts are required to establish the robustness and reliability of the proposed screening method. However, we believe that the identification of LS not only has the potential to prevent cancer-related morbidity and mortality but also facilitates continued progress in understanding the primary prevention of cancer.

## Data Availability

All data produced in the present study are available upon reasonable request to the authors.

## Ethnics approval

Our work was part of clinical study approved by the Ethical Committee of the Bratislava Self-Governing Region (Sabinovska ul.16, 820 05 Bratislava), on 30 November 2020 under the decision ID 09834/2020/HF. All patients in the study signed written informed consents consistent with the Helsinki declaration, which the ethics mentioned above committee approved.

## Conflict of interest statement

The authors Lucia Krasnicanova, Natalia Forgacova, and Vanda Repiska declare no conflicts of interest that could influence the work reported in this paper. Tatiana Sedlackova, Jaroslav Budis, Juraj Gazdarica, and Tomas Szemes are the employees of Geneton Ltd. which is involved in numerous research and development efforts dedicated to adapting new technologies to better understand genomic data and facilitate their implementation in effective and reliable patient care.

## Funding statement

This work was funded by the Operational program Integrated Infrastructure within the project: “Creation of nuclear herds of dairy cattle with a requirement for high health status through the use of genomic selection, innovative biotechnological methods, and optimal management of breeding” (NUKLEUS), ITMS code 313011V387, co-financed by the European Regional Development Fund.

## Data availability statement

All data produced in the present study are available upon reasonable request to the authors.

## Author contributions

LK and NF were responsible for the literature search and manuscript writing. LK and TSe were responsible for laboratory work. JG and JB performed data analysis. JG, JB and TSz were responsible for designing the study and supervising the work. TSz and VR conceived the idea of the project and performed proofreading of the manuscript. All of the authors confirm that they had full access to all the data in the study and take responsibility for the integrity of the data and the accuracy of the data analysis. All of the authors gave final approval of this version to be published and agreed to be accountable for all aspects of the work in ensuring that questions related to the accuracy or integrity of any part of the work are appropriately investigated and resolved.

## Notes

### Author Declarations

The Ethical Committee of the Bratislava Self-Governing Region (Sabinovska ul.16, 820 05 Bratislava), on 30 November 2020 under the decision ID 09834/2020/HF gave ethical approval for this work.

## References

1. Buglyó G, Styk J, Pös O, Csók Á, Repiska V, Soltész B, et al. Liquid Biopsy as a Source of Nucleic Acid Biomarkers in the Diagnosis and Management of Lynch Syndrome. Int J Mol Sci. 2022;23. doi:10.3390/ijms23084284

2. Styk J, Buglyó G, Pös O, Csók Á, Soltész B, Lukasz P, et al. Extracellular Nucleic Acids in the Diagnosis and Progression of Colorectal Cancer. Cancers. 2022;14. doi:10.3390/cancers14153712

3. Salikhanov I, Heinimann K, Chappuis P, Buerki N, Graffeo R, Heinzelmann V, et al. Swiss cost-effectiveness analysis of universal screening for Lynch syndrome of patients with colorectal cancer followed by cascade genetic testing of relatives. J Med Genet. 2022;59: 924–930.

4. Lepore Signorile M, Disciglio V, Di Carlo G, Pisani A, Simone C, Ingravallo G. From Genetics to Histomolecular Characterization: An Insight into Colorectal Carcinogenesis in Lynch Syndrome. Int J Mol Sci. 2021;22. doi:10.3390/ijms22136767

5. Duraturo F, Liccardo R, De Rosa M, Izzo P. Genetics, diagnosis and treatment of Lynch syndrome: Old lessons and current challenges. Oncol Lett. 2019;17: 3048–3054.

6. Trujillo-Rojas MA, Ayala-Madrigal M de la L, Gutiérrez-Angulo M, González-Mercado A, Moreno-Ortiz JM. Diagnosis of patients with Lynch syndrome lacking the Amsterdam II or Bethesda criteria. Hered Cancer Clin Pract. 2023;21: 21.

7. Cross DS, Rahm AK, Kauffman TL, Webster J, Le AQ, Spencer Feigelson H, et al. Underutilization of Lynch syndrome screening in a multisite study of patients with colorectal cancer. Genet Med. 2013;15: 933–940.

8. Umar A, Boland CR, Terdiman JP, Syngal S, de la Chapelle A, Rüschoff J, et al. Revised Bethesda Guidelines for hereditary nonpolyposis colorectal cancer (Lynch syndrome) and microsatellite instability. J Natl Cancer Inst. 2004;96: 261–268.

9. Adar T, Rodgers LH, Shannon KM, Yoshida M, Ma T, Mattia A, et al. Universal screening of both endometrial and colon cancers increases the detection of Lynch syndrome. Cancer. 2018;124: 3145–3153.

10. Moreira L, Balaguer F, Lindor N, de la Chapelle A, Hampel H, Aaltonen LA, et al. Identification of Lynch syndrome among patients with colorectal cancer. JAMA. 2012;308: 1555–1565.

11. Cohen SA, Pritchard CC, Jarvik GP. Lynch Syndrome: From Screening to Diagnosis to Treatment in the Era of Modern Molecular Oncology. Annu Rev Genomics Hum Genet. 2019;20: 293–307.

12. Styk J, Pös Z, Pös O, Radvanszky J, Turnova EH, Buglyó G, et al. Microsatellite instability assessment is instrumental for Predictive, Preventive and Personalised Medicine: status quo and outlook. EPMA J. 2023;14: 143–165.

13. Forgacova N, Gazdarica J, Budis J, Radvanszky J, Szemes T. Repurposing non-invasive prenatal testing data: Population study of single nucleotide variants associated with colorectal cancer and Lynch syndrome. Oncol Lett. 2021;22: 779.

14. Kiyozumi Y, Matsubayashi H, Horiuchi Y, Higashigawa S, Oishi T, Abe M, et al. Germline mismatch repair gene variants analyzed by universal sequencing in Japanese cancer patients. Cancer Med. 2019;8: 5534–5543.

15. Lynch HT, Lynch PM, Lanspa SJ, Snyder CL, Lynch JF, Boland CR. Review of the Lynch syndrome: history, molecular genetics, screening, differential diagnosis, and medicolegal ramifications. Clin Genet. 2009;76: 1–18.

16. Yamamoto H, Imai K. An updated review of microsatellite instability in the era of next-generation sequencing and precision medicine. Semin Oncol. 2019;46: 261–270.

17. Markow M, Chen W, Frankel WL. Immunohistochemical Pitfalls: Common Mistakes in the Evaluation of Lynch Syndrome. Surg Pathol Clin. 2017;10: 977–1007.

18. Bolger AM, Lohse M, Usadel B. Trimmomatic: a flexible trimmer for Illumina sequence data. Bioinformatics. 2014;30: 2114–2120.

19. Li H, Handsaker B, Wysoker A, Fennell T, Ruan J, Homer N, et al. The Sequence Alignment/Map format and SAMtools. Bioinformatics. 2009;25: 2078–2079.

20. Danecek P, Bonfield JK, Liddle J, Marshall J, Ohan V, Pollard MO, et al. Twelve years of SAMtools and BCFtools. Gigascience. 2021;10. doi:10.1093/gigascience/giab008

21. Budiš J, Krampl W, Kucharík M, Hekel R, Goga A, Sitarčík J, et al. SnakeLines: integrated set of computational pipelines for sequencing reads. J Integr Bioinform. 2023. doi:10.1515/jib-2022-0059

22. Siraj AK, Prabhakaran S, Bavi P, Bu R, Beg S, Hazmi MA, et al. Prevalence of Lynch syndrome in a Middle Eastern population with colorectal cancer. Cancer. 2015;121: 1762–1771.

23. Yurgelun MB, Hampel H. Recent Advances in Lynch Syndrome: Diagnosis, Treatment, and Cancer Prevention. Am Soc Clin Oncol Educ Book. 2018;38: 101–109.

24. Bhattacharya P, McHugh TW. Lynch Syndrome. StatPearls. Treasure Island (FL): StatPearls Publishing; 2023.

