## Supplementary material for "Optimizing mixed sample analysis as a step to comprehensive desease screening: a pilot study"

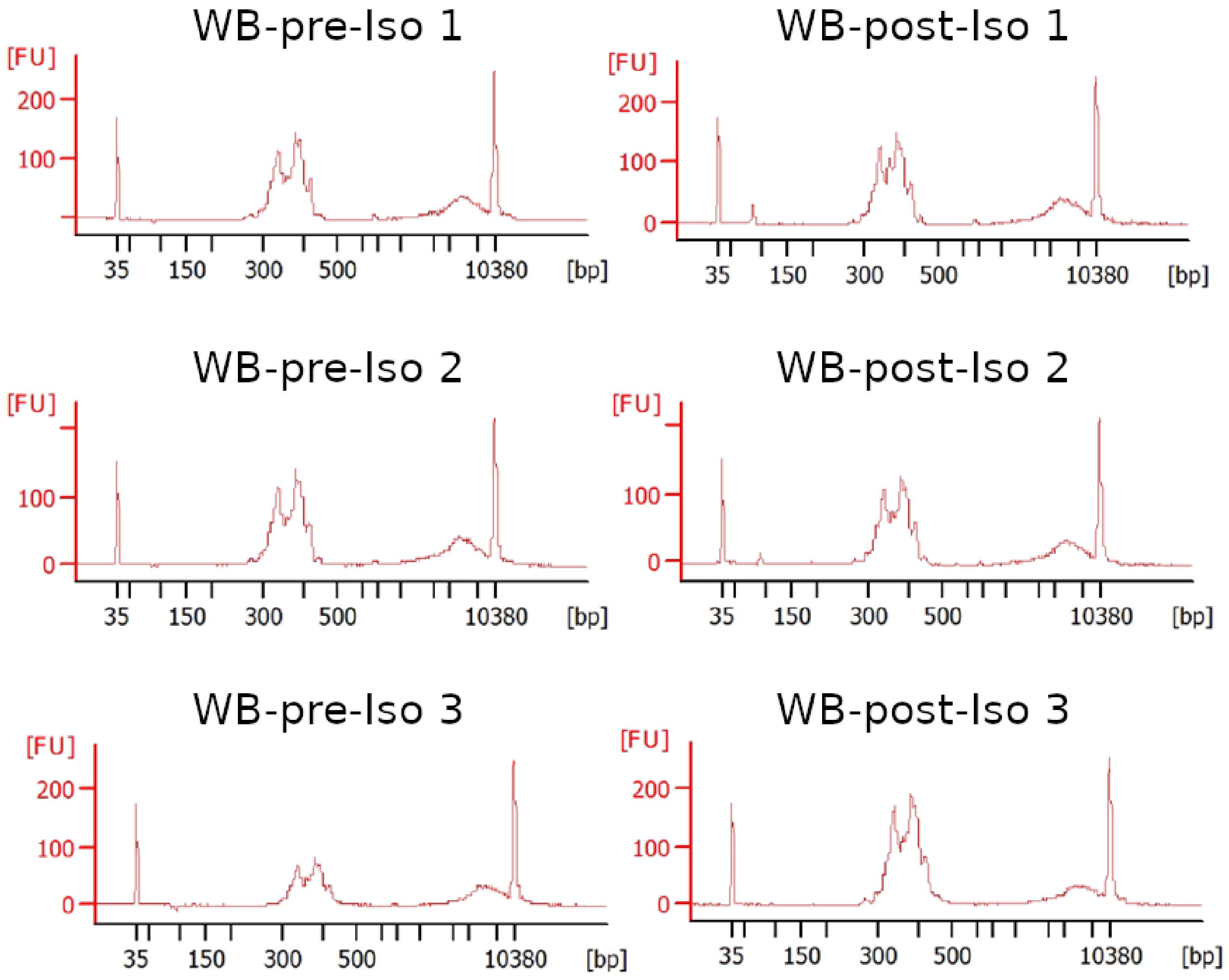


**Supplementary figure 1.** Quality analysis of the generated CleanPlex® Lynch Syndrome Panel (Paragon Genomics) DNA library using the Agilent 2100 Bioanalyzer for WB approaches.

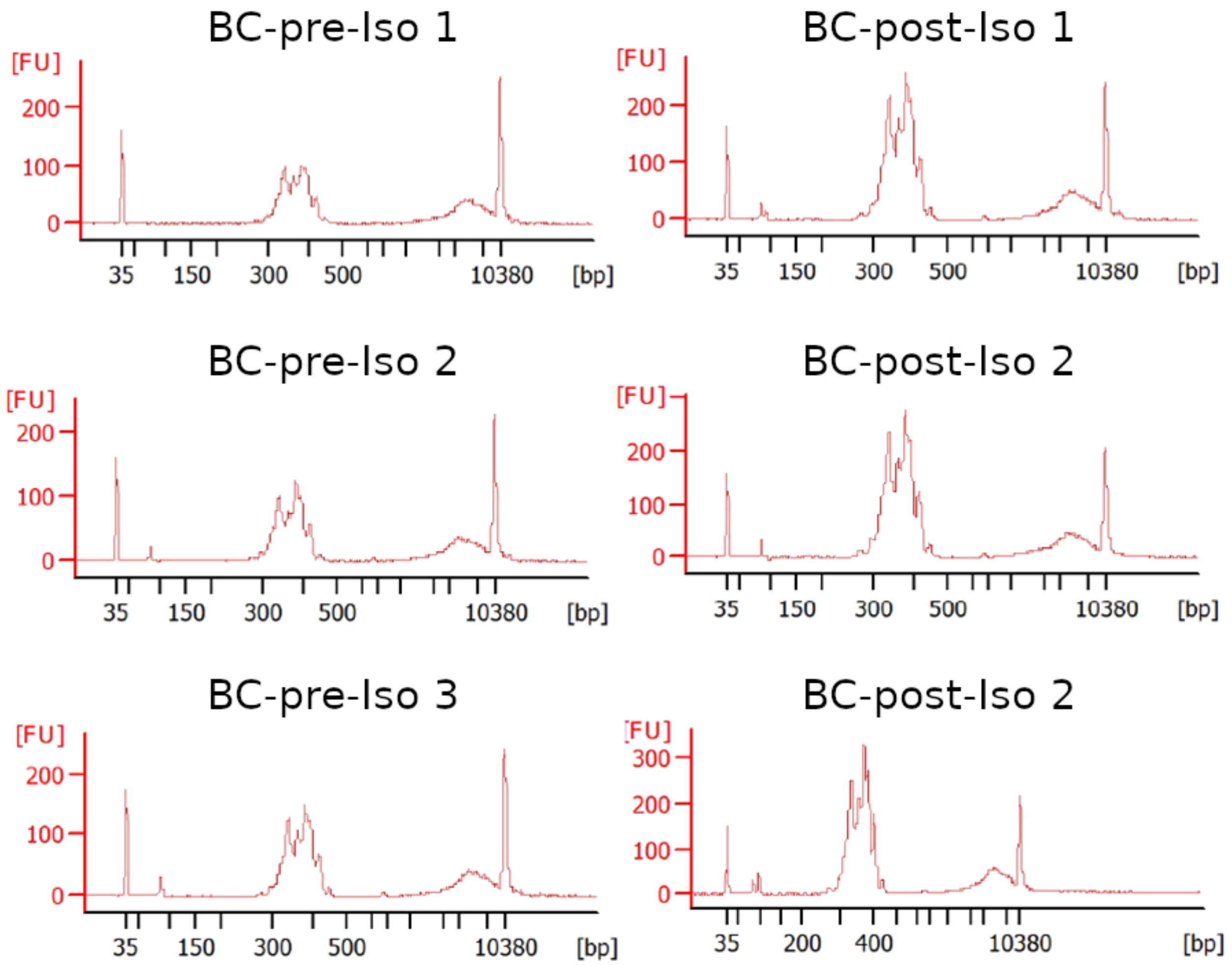


**Supplementary figure 2.** Quality analysis of the constructed CleanPlex® Lynch Syndrome Panel (Paragon Genomics) DNA library using the Agilent 2100 Bioanalyzer for BC approaches.

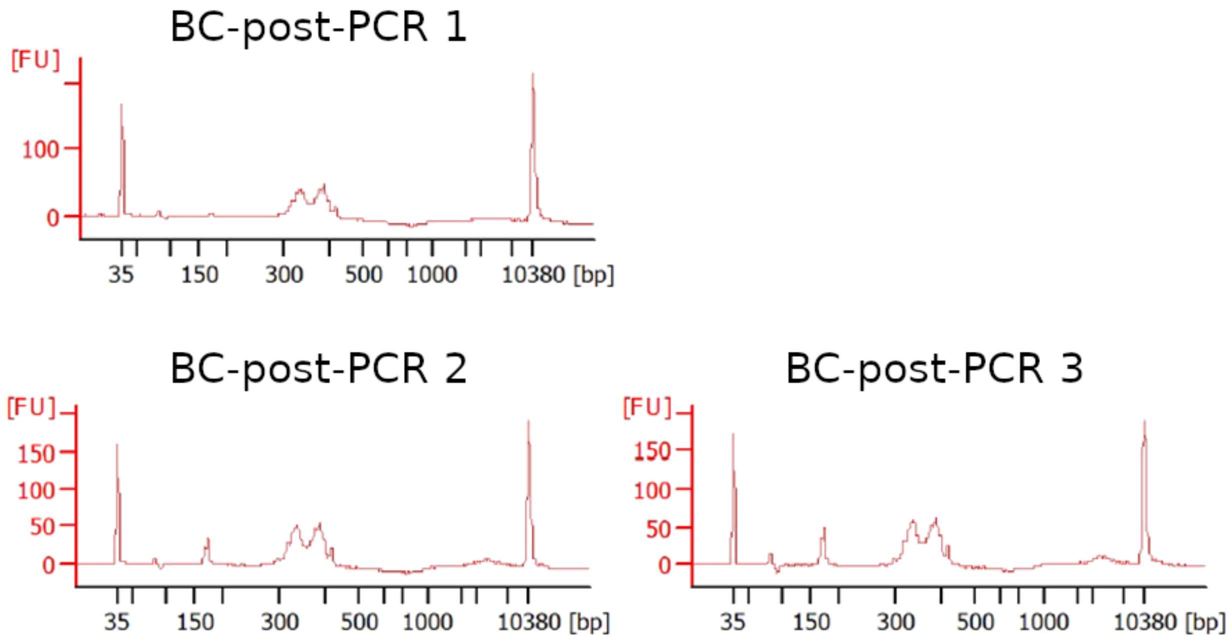


**Supplementary figure 3.** Quality analysis of the generated CleanPlex® Lynch Syndrome Panel (Paragon Genomics) DNA library using the Agilent 2100 Bioanalyzer for PCR amplification approach.

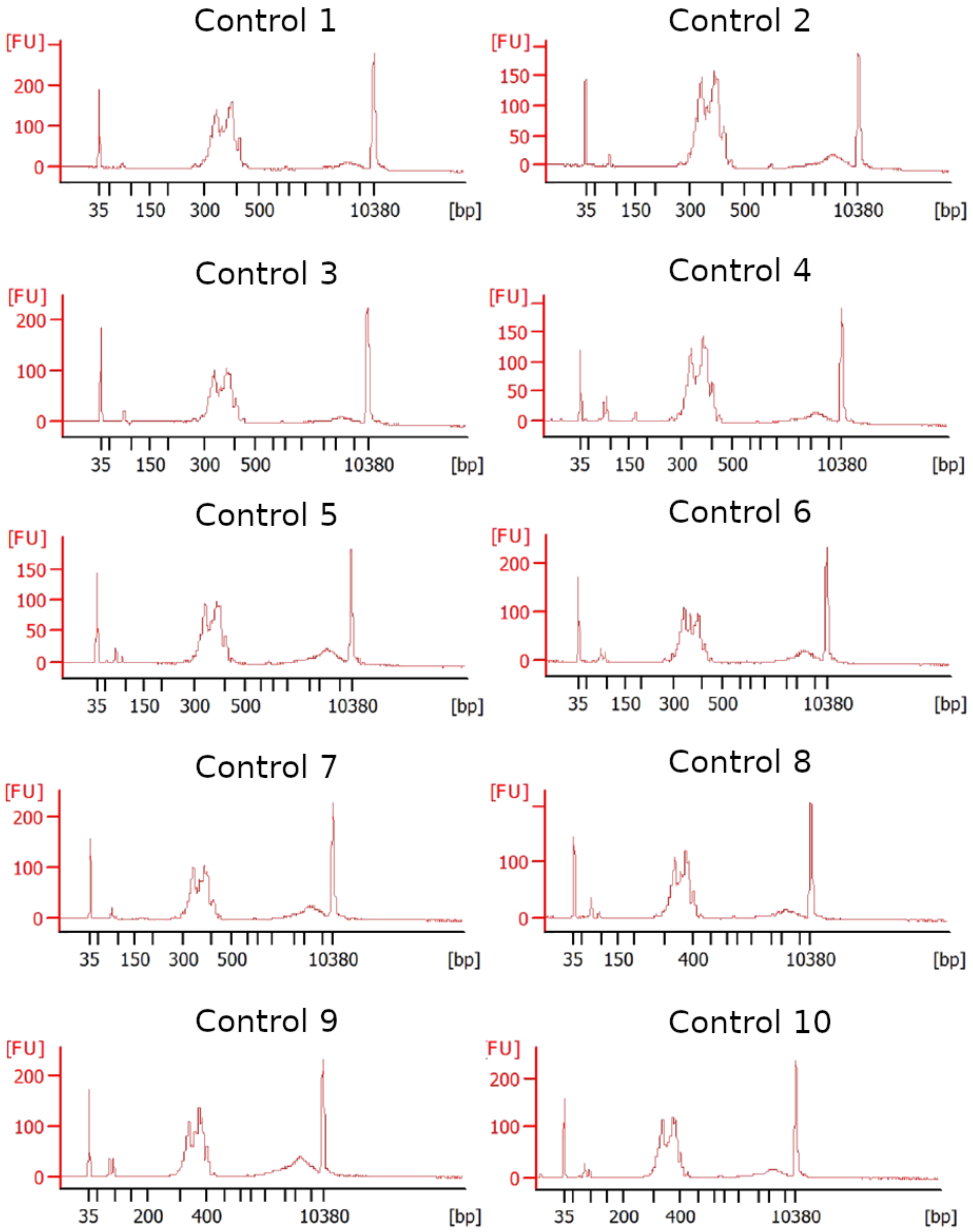


**Supplementary figure 4.** Quality analysis of the generated CleanPlex® Lynch Syndrome Panel (Paragon Genomics) DNA library using Agilent 2100 Bioanalyzer of each sample separately.
